# Managing anginal chest pain in primary care: A qualitative analysis with perspectives from general practitioners

**DOI:** 10.1101/2025.10.15.25337722

**Authors:** Kim MG van Bergen, Moniek Y Koopman, Birgitt Klerkx, Gigja Vossen, Rozemarijn Vliegenthart, Geert-Jan Dinant, Robert TA Willemsen, Eefje GPM de Bont, Jochen WL Cals

## Abstract

**Aim:** The aim of this study was to gain insight into general practitioners’ (GPs’) modus operandi, by investigating the experiences of GPs regarding the assessment of anginal chest pain.

**Introduction:** Assessing anginal chest pain in primary care is hampered by a presentation that is often different from text-books and by limited diagnostic tools.

**Methods:** We conducted a qualitative, semi-structured interview-based study among GPs in the Netherlands regarding diagnosis, classification, referral, treatment and guideline adherence in anginal chest pain.

**Findings & conclusions:** We interviewed 16 GPs. They expressed challenges in classification of chest pain, including determining its typicality and distinguishing stable from unstable chest pain. These uncertainties, combined with the limited direct access to diagnostics in primary care, lead to diversity in diagnostics approach, treatment and referrals of patients with anginal chest pain, next to doubts regarding the guideline’s referral criteria. This lead GPs to combine guideline adherence with personal experience and intuition in dealing with anginal chest pain. GPs aim to balance avoiding unnecessary referrals with minimizing missing chronic coronary syndrome diagnoses, resulting in diverse anginal chest pain management strategies in primary care. GPs emphasize the need for improvements in diagnostics and advanced reassurance methods to safely and effectively exclude severe disease.

## Introduction

### Anginal chest pain in primary care

In primary care, chest pain is a common complaint with an incidence of 1.5 per 1000 patients per year in the Netherlands (1). The differential diagnosis is broad and dominated by non-life threatening causes, but the possibility of severe underlying disease cannot be neglected, as a life threatening cause is present in 3 to 8.4% of cases (2, 3). Therefore, prompt diagnosis and treatment are essential (4).

Anginal chest pain (ACP) is the main manifestation of chronic coronary syndrome (CCS), incorporating atherosclerotic large vessel disease or myocardial microvascular dysfunction (small vessel disease). Stable ACP is further categorised based on the presence of three core symptoms: retrosternal pain, provocation by exercise or stress, and relief at rest or with nitrates (4, 5). If all are present, it is referred to as typical angina pectoris (AP); when two are present, it is considered atypical AP; and when one or none are present, it is classified as non-anginal chest pain or non-specific thoracic complaints (4).

### Challenges in the management of anginal chest pain

If (a)typical AP is present, the probability of CCS is present and referral to the cardiologist is indicated according to both the updated Dutch primary care guideline on ACP and the European Cardiology guideline, whereas it is recommended not to refer patients with non-anginal chest pain because of the low probability of CCS (4, 5).

However, typical AP, the easiest recognizable presentation, is present in only 13-16% of patients suspected of CCS (6, 7). Furthermore, the presentation of chest pain is increasingly less classical, making it difficult to assess typicality of symptoms (8). As a consequence, the challenge in the assessment of complaints leads to unnecessary diagnostics, treatment and/or referrals, which is accompanied by high costs (1, 9, 10). Certainly, although a wide variety of diagnostic tests is performed in primary care, such as electrocardiograms (ECGs), chest X-rays and Hemoglobin tests, the revised guidelines no longer recommend performing exercise ECGs, reflecting changes in best practices (3, 5, 10).

### Aim

To date, no studies have explored the experiences or perspectives of general practitioners (GPs) on managing anginal chest pain in primary care. The majority of studies mainly focus on describing diagnostic behaviour and accuracy of GPs (3, 11, 12). The aim of this study is therefore to gain more insight into the thoughts of GPs as well as possible barriers or doubts they experience when assessing ACP in daily practice.

## Methods

### Research approach

In a qualitative, interview-based study among GPs in the Netherlands, we conducted interpersonal, unmediated, semi-structured interviews with open-ended questions. These were conducted between November 2020 and April 2021, either in person at participants’ practices or via secured Zoom meetings when COVID-19 restrictions limited face-to-face contact.

### Sampling, data saturation

We alerted Dutch GPs through a variance of communication channels of the department of general practice (‘purposive sampling’), such as e-mail, via an article in a magazine and by using social media, with all media together having a reach of 1884 GPs. We purposely focused on Dutch GPs with at least two years of working experience, to guarantee their clinical experience. GPs with a specialization in cardiovascular diseases were excluded to avoid potential bias in perspectives. Baseline characteristics of the study population included age, gender, years of experience as a GP, type of practice (single/duo/group), position (employee/practice owner), area (urban/rural) and field of interest. We requested participation confirmation via email or telephone. Interviews continued until data saturation and inductive thematic saturation was reached, defined as the lack of obtainment of new theoretical insights, new themes or new codes emerging in three consecutive interviews (13, 14).

### Interview

#### Box 1.

##### Interview Topic guide

###### Leading subjects

1. recent case with stable angina pectoris in the GPs practice
2. and attitude towards the Dutch GP guidelines in general
3. and attitude towards the revised Dutch GP guideline ACP
4. regarding the classification of chest pain symptoms
5. regarding non-anginal chest pain
6. and attitude towards diagnostics, treatment and referrals for patients with ACP
7. for additions, and themes GPs had missed during the interviews

The interview (see Appendix 1) consisted of seven main subjects to structure the interview, as described in box 1. During the study, questions were open for adjustment, following the topics initiated by GPs themselves. The research team consisted of two researchers and two medical trainees. Both researchers are medical doctors. After a pilot interview by one researcher and two medical trainees (RW, GV and BK), the medical trainees (GV and BK) conducted all interviews face-to-face, with audio recording of each interview. One researcher anonymously transcribed verbatim all audio interview recordings and sent to each individual study participant for possible additions, corrections and final approval. All participating GPs signed for informed consent before conducting the interviews. There were no prior relationships or personal interactions between the researchers and the participants in this study.

### Data analysis and trustworthiness

We conducted a conventional analysis, focusing on interview texts as the primary data (15). After development of a codebook, we analysed all transcripts by inductive coding with an open procedure. Two researchers coded the first three transcripts independently to obtain agreement on the method of coding. Thereafter, one researcher finished coding of the remaining interviews and discussed ambiguities or doubts with the research team. We used NVivo 12 software for encoding. Codes were classified into several overarching descriptive themes in close dialogue with the research team (BK, GJD, RW), followed by a thematic analysis of all data by RW, GJD (both experienced GPs) and BK. Three researchers enhanced triangulation by reviewing, discussing and interpreting the data, ultimately reaching consensus. Data triangulation incorporated the diverse backgrounds of GPs, including variations in experience, practice type and geographic location. While fieldnotes were not employed, the analysis relied on comprehensive transcripts of the interviews. Study reporting aligns with the Consolidated criteria for Reporting Qualitative Research (COREQ) criteria (16).

### Ethical approval

We informed all research participants in advance about the research objectives, recordings and transcripts of the interviews. The Medical Ethical Review Committee (METC) of Maastricht University reviewed and approved the study.

## Results

We performed 16 interviews before complete data saturation was accomplished (13 initial interviews plus three additional interviews to confirm saturation). All GPs agreed on participation and completed informed consent. The duration of the interviews varied between 30-40 minutes. Table 1 illustrates the baseline characteristics of the 16 GPs participating in the study. The specific areas of interest among GPs showed great diversity, with only one GP indicating a personal interest in cardiology. Figure 1 summarizes the five research themes, and the main findings based on the semi-structured interviews.

**Figure 1.**
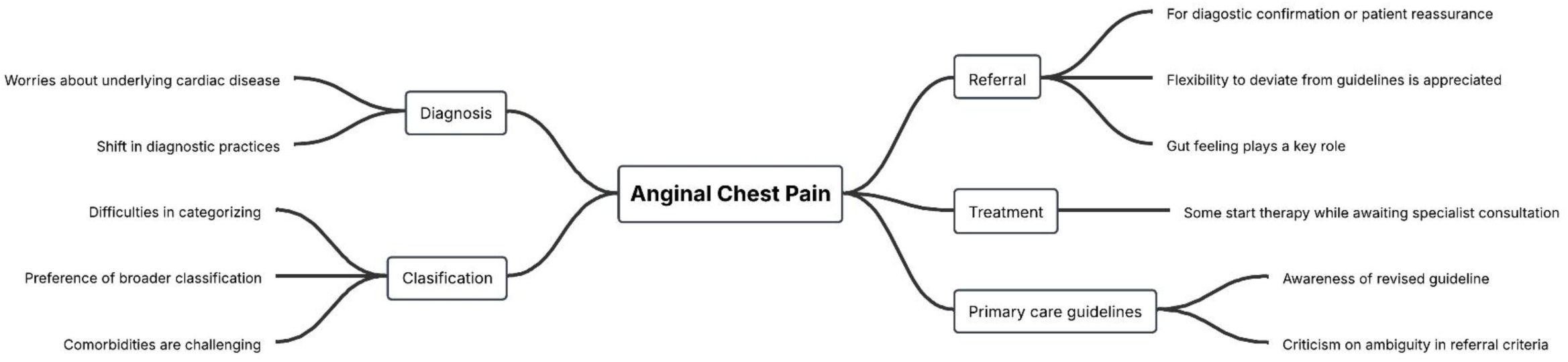
Main findings on the five research themes

**Table 1.**
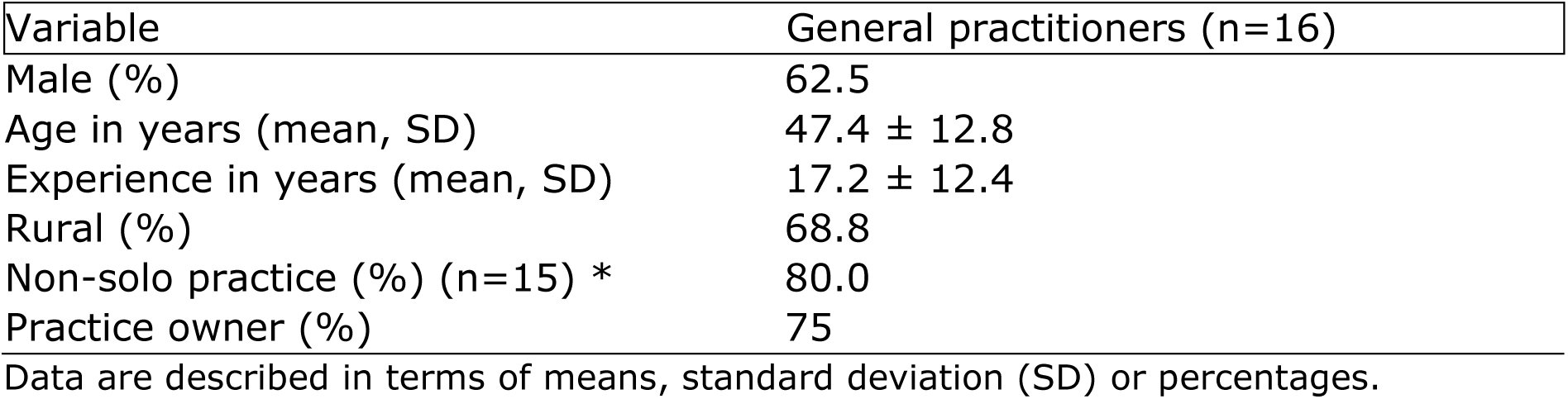
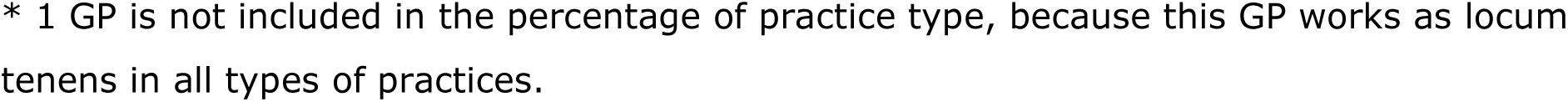
Baseline characteristics GPs (n=16).

| Variable | General practitioners (n=16) |
| --- | --- |
| Male (%) | 62.5 |
| Age in years (mean, SD) | 47.4 ± 12.8 |
| Experience in years (mean, SD) | 17.2 ± 12.4 |
| Rural (%) | 68.8 |
| Non-solo practice (%) (n=15) * | 80.0 |
| Practice owner (%) | 75 |

Data are described in terms of means, standard deviation (SD) or percentages.
\* 1 GP is not included in the percentage of practice type, because this GP works as locum tenens in all types of practices.

### Theme 1: diagnosis

The GPs in this study report that the incidence of new-onset ACP in primary care is low, possibly due to the adequate cardiovascular risk management programmes in The Netherlands.

While there is consensus about the infrequency of new-onset stable angina, GPs struggle with distinguishing between stable and unstable angina, often perceiving the distinction as a grey area.

GP 14: ‘(..)If it is someone who has never had cardiac complaints and suddenly experiences exercise-bound pain, even if it is always with the same exertion, but it only exists for one week or one and a half week, then I believe it is unstable (..)’

Regarding the management of ACP, GPs express that both patients and clinicians still experience significant worries about underlying cardiac disease.

GP 1: ‘With stable angina pectoris, I prefer it that they also have been seen by the cardiologist. Sometimes you get reassurance, and that works well here in the region.’

The study highlights varying perspectives on the diagnostic limitations in primary care. While some GPs prefer to refer patients with suspected ACP directly to a cardiologist, others express confidence in conducting additional diagnostic tests themselves in primary care. Although advised otherwise in the revised guideline, several GPs indicated they will continue ordering exercise ECGs as a reassurance tool, particularly for patients with a low likelihood of cardiac conditions, since the test remains directly accessible in their region. Other GPs expressed concerns about the reliability of the test that in their opinion fails to definitively rule out or confirm the presence of a cardiac condition. At the same time, these GPs indicate they require time to integrate the updated guidelines into their practice.

The shift in diagnostic practices is viewed by several GPs as a change in the wrong direction, as it may lead to an increase in referrals to secondary care, due to the limited access to indicated diagnostic resources in primary care. In response to the altered primary care guideline, GPs indicate the need for more feedback on referral decisions and insights into the diagnostic steps followed by cardiologists.

GP 2: ‘(..) What I would like to hear back is when is it indeed useful to refer someone for outpatient care and within what period. And what steps does a cardiologist follow. This also gives us insight into whether it is useful to refer.’

Finally, GPs suggest the introduction of new diagnostic tools to help reduce unnecessary referrals and tests. Options mentioned include home-based ECG, direct-access echocardiography, GP access to CT calcium score, access to one-stop outpatient consultations by the cardiologist or interdisciplinary electronic consultation with the cardiologist, and GP access to validated point of care tests and a decision tree or score list.

### Theme 2: classification

GPs report difficulties in categorizing chest pain as (a)typical AP or non-anginal chest pain (or non-specific thoracic pain). Some are viewing atypical AP and non-anginal chest pain as a diagnostic “grey area”. Others acknowledge uncertainty in using these terms correctly.

The Dutch primary care guideline classification is perceived as artificial, as AP presentation is seen as a continuum rather than three well-defined categories. In reality, more factors should be taken into consideration. GPs fear missing CCS if only the three core symptoms are considered.

GP 12: ‘I think it’s quite artificial actually. (…) In practice there are of course other things involved. What history does someone have? Does someone smoke? Does someone come easily to the consultation hour, or does he never come? And what kind of gut feeling do you have? (…) If I think back for a moment, I can tell a few cases where I had a bit of a bad feeling, and indeed heart complaints were found. And they would not have been referred according to these criteria.’

Some GPs use the criteria to define (a)typical AP or non-anginal chest pain. The majority of GPs prefer a broad classification, distinguishing cardiac from non-cardiac conditions rather than strictly applying guideline-based categories.

GP 3: ‘I never write down ‘typical’ and ‘atypical’ AP as such anyway. I do write down ‘non-specific thoracic pain’ complaints and ‘AP’. So, then it is just one of those two. It is something or it is nothing. (…)’

Moreover, diagnosing anginal chest pain in patients with comorbidities (e.g., COPD, psychiatric conditions, or somatization) is particularly challenging, as overlapping symptoms complicate assessment and may limit the patient’s capacity to exert themselves. Besides, diabetic patients are often referred earlier due to their higher cardiovascular risk, while women undergo more diagnostic testing and referrals due to atypical symptom presentation, such as fatigue or epigastric pain.

Despite primary care guideline recommendations, GPs refer patients with non-anginal chest pain when uncertainty remains about the absence of an underlying cardiac condition.

‘GP 5: (…)I once learned that you don’t get a guideline of keeping people at home, so when in doubt, I prefer to go for the diagnostics. (…) Eventually you risk harming your patients and yourself if you don’t and something comes to light anyway.’

### Theme 3: referral

Most GPs in this study refer patients with suspected ACP to secondary care, primarily for diagnostic confirmation or patient reassurance. Moreover, an increasing tendency to refer over the years is seen, due to advancements in cardiology.

GPs generally find the primary care guideline’s recommendation to refer patients with (a)typical AP, while not referring those with clear non-anginal chest pain, to be feasible, as with this approach probably less severe cases will be missed and more patients will be referred to secondary care. However, they appreciate the flexibility to deviate in specific cases, such as elderly patients with comorbidities or those unwilling to be referred.

Despite this, GPs occasionally remain uncertain about not referring patients with supposedly non-anginal chest pain due to diagnostic limitations.

‘GP 5: (…) From time to time there are people who you refer immediately anyway, and on the other side there are also people who make you think: this is nothing at all and then it turns out to be a cardiac story after six months. So, there is a grey area in it. And that … well, that is difficult to include in the guideline to prevent you from getting overdiagnosis or underdiagnosis.’

Concerns about missing severe cases influence referral decisions. Some GPs refer patients with non-anginal chest pain if uncertainty persists, particularly in cases of positive family history, significant risk factors (e.g., diabetes), or worried patients.

While some GPs see the revised guideline as a step forward—reducing missed cases by increasing referrals—others worry about overwhelming secondary care or still failing to detect severe cases.

Gut feeling plays a key role in decision making, especially in non-anginal chest pain cases. GPs cite diagnostic uncertainty, unexplained symptoms, or repeated visits as triggers for concern.

GP 8: ‘(…) If I cannot explain it properly, then it also depends a bit on the sex, age and history… if I don’t have a good explanation for it, then the gut feeling does come up with me.’

A minority of the GPs refers immediately without prior diagnostic testing. GPs advocate for incorporating shared decision-making, patient context, and risk factors—especially sex differences—into the guideline. Some GPs indicate they possibly refer women sooner due to their less typical symptom presentation.

A GP proposed that establishing intermediate care centres could mitigate the overflow of referrals to secondary care, which has already been established in a region in the southern part of the Netherlands.

GP 10: ‘That is also why the intermediate care centre has arrived in that region. That [anonymous] region was at one point a somewhat unhealthy region, which overflowed a bit, and they capture many such referrals.’

### Theme 4: treatment

GPs report that pharmacological treatment for ACP is typically initiated by the cardiologist, after which patients return to primary care. Most GPs start some therapy while awaiting specialist consultation, whereas others wait for the cardiologist’s assessment before initiating treatment decisions.

GP 8: ‘(…) I start with a platelet aggregation inhibitor, beta blocker and a nitro-glycerine spray. With the idea that a lipid spectrum and possibly the coronaries will eventually be addressed by the cardiologist.’

A minority of GPs manage ACP independently without referral, particularly in patients with significant comorbidities, advanced age, or limited quality of life, considering patient preferences.

### Theme 5: primary care guidelines in general and concerning ACP

Most GPs are aware of the revised Dutch primary care guideline on ACP and generally adhere to its recommendations. As these guidelines are already familiar, GPs consult them infrequently (0–2 times per year). While they acknowledge the value of evidence-based, standardized recommendations, some consciously deviate in specific cases.

The revised primary care guideline on ACP is praised for its clarity but criticized for ambiguity in referral criteria, particularly in cases of non-anginal chest pain. GPs perceive conflicting messages: a warning to avoid missing severe cases alongside advice to limit unnecessary referrals. Some GPs mention the fact that the Dutch guideline regarding ACP focuses partially too much on numbers and to a lesser extent on the person-oriented care.

GP 12: ‘That pinches a bit. On the one hand, we are extremely warned about it. On the other hand, it is said: be restrictive with your referrals. There is, of course, a field of tension.’

## Discussion

### General discussion

In this study, five overarching themes were defined. First, concerning the **diagnosis** of ACP, both patients and GPs often associate anginal chest pain with severe disease and as a consequence GPs experience diagnostic uncertainty. Indeed, underlying life-threatening causes, such as acute coronary syndrome, pulmonary embolism or aortic aneurysm, are found in 8.4% of patients presenting with chest pain in primary care (3, 8). On the other hand, a majority of patients can be reassured but distinction of these patients from patients with severe cardiac disease is challenging (17). The uncertainty experienced by GPs is further emphasized in another study that found sensitivity and specificity of GPs for CCS, to be 69% (95% CI: 62%-75%) and 89% (95% CI: 87%-91%) respectively, highlighting areas for improvement (11).

Comorbidities (e.g. COPD, psychiatric disorders, type 2 diabetes) are considered to complicate a clear diagnosis by increasing the atypicality of symptoms and comprising symptoms equal to CCS. Participating GPs mention that they more frequently refer women for diagnostic testing or consultation by a cardiologist, due to fears of missing a diagnosis as a consequence of less typical presentations. Possibly, this is related to campaigns raising GP awareness of cardiac ischemia in women. On the other hand, literature and the Dutch guideline highlight more similarities than differences in chest pain presentation between sexes (5, 18–20).

GPs often classify new-onset chest pain persisting for several weeks as unstable AP. However, in the guideline, type of pain and effort dependency are the leading criteria to distinguish stable from unstable chest pain, not symptom duration (4, 5). Most GPs align with the Dutch and European guidelines advising against the use of exercise ECGs to confirm or rule out a cardiac cause in patients presenting with chest pain (4, 5, 21).

Nevertheless, some GPs continue to employ it as a standalone test to exclude CCS in cases with a presumed low pretest probability, primarily to reassure the patient and themselves.

GPs in this study expressed a desire for advanced diagnostic tools in primary care, such as coronary CT scans, point-of-care tests or clinical decision rules, corresponding to earlier studies (12, 21, 22). On the other hand, GPs emphasize the value of reassuring patients through detailed explanations or organized multidisciplinary care (aided by physical therapists and psychologists), which could reduce expensive and unnecessary diagnostic procedures (1, 10). Finally, GPs stated that a better understanding of the cardiologist’s workup could enhance their ability to inform patients effectively, in line with literature highlighting opportunities to improve communication between GPs and cardiologists (23).

Secondly, in the **classification** of ACP, GPs do not strictly adhere to the three criteria (type of pain, worsening on exertion, relief with rest or sublingual nitro-glycerine) as stated in the Dutch primary care guideline. Both our study and others confirm that additional factors are taken into account, such as history of cardiovascular disease, age, cardiovascular risk factors, comorbidities, context, prior knowledge of the patient, the GP’s gut feeling and the patient’s belief in a cardiac origin (4, 17, 24). GPs use the terms typical AP, atypical AP and non-anginal chest pain interchangeably and admitted the definitions were not ‘on top of their mind’. They partly attribute this to the commonly used International Classification of Primary Care (ICPC) coding system, where AP can only be registered as stable or unstable. The European guideline identifies these terms as potentially misleading and recommends estimating the clinical likelihood of CCS based on a broader combination of symptom characteristics and risk factors (4). Moreover, the American guideline highlights that atypical chest pain is often used to describe non-anginal symptoms (17). A phenomenon confirmed by most of our participating GPs.

Furthermore, they tend to adopt an approximate binary approach, classifying thoracic complaints either suspected cardiac or non-cardiac in origin. The American guideline recommends clearer classifications such as ‘cardiac’, ‘non-cardiac’ and ‘possible cardiac’. It could be considered to re-evaluate the descriptions provided within the Dutch guidelines in line with the more extended criteria in the European and/or the more practice-based judgement in the American guidelines.

However, GPs appear to implicitly implement the three criteria of typicality in assessing patients with ACP, integrating them in their interpretation of complaints, also addressing their gut feeling. Previous European studies have shown that in primary care, gut feeling arises from GP’s prior knowledge and experience, and on pattern recognition. It aids in differentiating urgent from non-urgent conditions, especially in cases of uncertainty (25–27). In our study, GPs mention gut feeling as a key tool in evaluating chest pain complaints, incorporating observations such as incongruity with patient context, absence of reassuring alternative diagnoses, and repeated patient visits after initial reassurance.

Third, GPs **refer** the majority of patients suspected of (a)typical AP, although opinions on the current Dutch primary care guideline are mixed. While some believe the changes improve referral accuracy and reduce missed diagnoses of CCS, others express concerns about overlooking life-threatening conditions. Despite guidelines discouraging referrals for non-anginal chest pain to cardiologists, GPs frequently do so to rule out cardiac ischemia, prioritizing reassurance over guideline adherence (5). Moreover, GPs highlight the need for more guidance on shared decision-making in the Dutch primary care guideline.

Fourth, a great diversity is seen in the **treatment** in patients with ACP. Some GPs initiate therapy themselves in patients they refer, while others await the advice of the consulted cardiologist. Variation in the type of medication initiated was seen, although the guideline clearly states the recommendation to start a platelet aggregation inhibitor, a statin and sublingual application of nitro-glycerine when needed (5). Regarding initiation of treatment in ACP, GPs emphasize that the recommendations are occasionally not followed in practice, which is in accordance with findings in a previous study (23).

Fifth, GPs embrace the Dutch primary care **guidelines**, including the ACP guideline, as a key framework to guide their daily work. Knowledge regarding the management of ACP was adequate among GPs in our study. They generally adhere to the recommendation on typicality of complaints, treatment and referrals, though sometimes they consciously deviate from the guideline, especially for older patients, those with comorbidities, or in response to the patient’s wishes. Awareness of the revised ACP recommendations varies, sometimes leading to only gradual replacement of old patterns. GPs highlight that the guideline lacks clarity on reassuring patients with non-anginal chest pain. As previously mentioned, this creates tension between avoiding missed diagnoses of cardiac ischemia and minimizing unnecessary referrals. They note that the ACP guideline heavily focuses on evidence-based recommendations, often overlooking clinical experience, gut feeling and patient context (3, 8, 27).

### Strengths and limitations

In this investigation, GPs perspectives were leading. Another strength of this study is the number of three additional interviews to confirm saturation, which enhances the reliability and depth of the insights into ACP management in primary care. However, some limitations were noted. First, the voluntary participation may have led to an overrepresentation of GPs with a specific interest in the topic, though this is unlikely to bias findings significantly as only one GP mentioned a particular interest in cardiology.

Second, some participants may have reviewed the revised primary care guideline before the interview, enhancing more desirable answers in this study. However, open-ended questions in these in-depth interviews encouraged honest reflections on personal experiences and beliefs, rather than testing guideline knowledge, which was not the aim of this study. Third, most participants were from the South of the Netherlands, but uniform national primary care guidelines ensure generalizability on the management of ACP. Fourth, regarding trustworthiness, we deliberately chose not to conduct a member check due to practical constraints and the potential influence of social desirability on participant responses. Finally, the study only considered GPs’ perspectives, excluding those of patients or cardiologists.

## Implications and conclusion

This interview study explored GPs’ perspectives on managing anginal chest pain symptoms in primary care. Several findings are noteworthy. First, the Dutch primary care guideline’s division into (a)typical AP and non-anginal chest pain is deemed arbitrary by GPs, who prefer comprehensive evaluations based on classical history-taking, patient context, gut feeling and previous experience to adequately deal with the insecurity experienced when assessing anginal chest pain. Future research is required to determine which aspects of GPs’ approaches should be preserved and improved. Second, the efficiency and safety of referring patients with (a)typical AP and not referring patients with non-anginal chest pain is doubted by GPs. Third, duration of complaints is seen as more critical than exercise-related symptoms for defining (un)stable chest pain, contrary to guidelines. Fourth, GPs associate female sex with atypical cardiac presentations, sometimes influencing their management and prompting lower thresholds for tests or referral, despite limited evidence. Fifth, GPs continue using exercise ECGs as stand-alone tests for excluding CCS, contrary to guideline recommendations. Lastly, advances in diagnostics and reassurance methods without additional diagnostic tests are desired by GPs.

## Data Availability

All data produced in the present study are available upon reasonable request to the authors

## Acknowledgements

None

## Authors’ Contribution

KB: Visualization, Writing – original draft. MK: Writing – review & editing. BK: Conceptualization, Formal analysis, Investigation, Methodology. GV: Formal analysis, Investigation. RV: Writing – review & editing. GJD: Validation, Supervision, Conceptualization. RW: Writing – review & editing, Validation, Supervision, Conceptualization. EB: Writing - review & editing, Validation, Supervision. JC: Writing – review & editing, Supervision

## Ethical standards

The authors assert that all procedures contributing to this work comply with the ethical standards of the relevant national and institutional guidelines on human experimentation (Maastricht University) and with the Helsinki Declaration of 1975, as revised in 2008. Written informed consent was obtained from all participants.

